# ClinGen Variant Curation Interface: A Variant Classification Platform for the Application of Evidence Criteria from ACMG/AMP Guidelines

**DOI:** 10.1101/2021.02.12.21251663

**Authors:** Christine G. Preston, Matt W. Wright, Rao Madhavrao, Steven M. Harrison, Jennifer L. Goldstein, Xi Luo, Hannah Wand, Bryan Wulf, Gloria Cheung, Mark E. Mandell, Howard Tong, Shaung Cheng, Michael A. Iacocca, Arturo Lopez Pineda, Alice B. Popejoy, Karen Dalton, Jimmy Zhen, Selina S. Dwight, Lawrence Babb, Marina DiStefano, Julianne M. O’Daniel, Kristy Lee, Erin R. Riggs, Diane B. Zastrow, Jessica L. Mester, Deborah I. Ritter, Ronak Y. Patel, Sai Lakshmi Subramanian, Aleks Milosavljevic, Jonathan S. Berg, Heidi L. Rehm, Sharon E. Plon, J. Michael Cherry, Carlos D. Bustamante, Helio A. Costa, on behalf of the Clinical Genome (ClinGen) Resource

## Abstract

**Background:** Identification of clinically significant genetic alterations involved in human disease has been dramatically accelerated by developments in next-generation sequencing technologies. However, the infrastructure and accessible comprehensive curation tools necessary for analyzing an individual patient genome and interpreting genetic variants to inform healthcare management have been lacking.

**Results:** Here we present the ClinGen Variant Curation Interface (VCI), a global open-source variant classification platform for supporting the application of evidence criteria and classification of variants based on the ACMG/AMP variant classification guidelines. The VCI is among a suite of tools developed by the NIH-funded Clinical Genome Resource (ClinGen) Consortium, and supports an FDA-recognized human variant curation process. Essential to this is the ability to enable collaboration and peer review across ClinGen Expert Panels supporting users in comprehensively identifying, annotating, and sharing relevant evidence while making variant pathogenicity assertions. To facilitate evidence-based improvements in human variant classification, the VCI is publicly available to the genomics community and is available at https://curation.clinicalgenome.org. Navigation workflows support users providing guidance to comprehensively apply the ACMG/AMP evidence criteria and document provenance for asserting variant classifications.

**Conclusion:** The VCI offers a central platform for clinical variant classification that fills a gap in the learning healthcare system, and facilitates widespread adoption of standards for clinical curation.

## Introduction

The application of genomics to precision medicine holds great promise for the implementation of tailored diagnostics, optimized patient care management, and personalized therapies in healthcare. The past decade has seen the development of technological and computational innovations to bring both DNA-sequencing methodologies and bioinformatic algorithms into routine standard-of-care for diagnostic medical genomics. While there has been broad consensus in terms of bioinformatic best practices, quality control metrics, and community adoption of variant calling and classification standards, substantial variability remains among variant curation tools and data sharing by health care providers, clinical diagnostic laboratories, and researchers.

Clinical interpretation of genomic sequencing data requires both the standardization of variant classification guidelines, as well as consistency in the workflow and evidence considered when determining the relationship between a variant and a disease phenotype. Since the 2015 release of the American College of Medical Genetics and Genomics / Association for Molecular Pathology (ACMG/AMP) guidelines^1^, the genomics community has substantially adopted^2^ the standards outlined by Richards *et al*. and the guidance provided by the Clinical Genome Resource (ClinGen) Consortium^3^ to aid in further refining and standardizing the evaluation of sequence variant pathogenicity^4–16^.

While the creation of these variant classification guidelines and standards is a central objective, there is also a significant unmet need for computational infrastructure and curation software to guide biocurators through these complex curation workflows. Here we present the ClinGen Variant Curation Interface (VCI), which is a comprehensive variant classification platform designed to support both individual and group classification in accordance with the ACMG/AMP guidelines. The VCI is intended to be a publicly available variant curation tool which programmatically guides users through a standard process for variant evidence classification and application of ACMG/AMP guidelines in a controlled workflow to enforce rigor and quality in variant classification (**Figure 1**).

**Figure 1:**
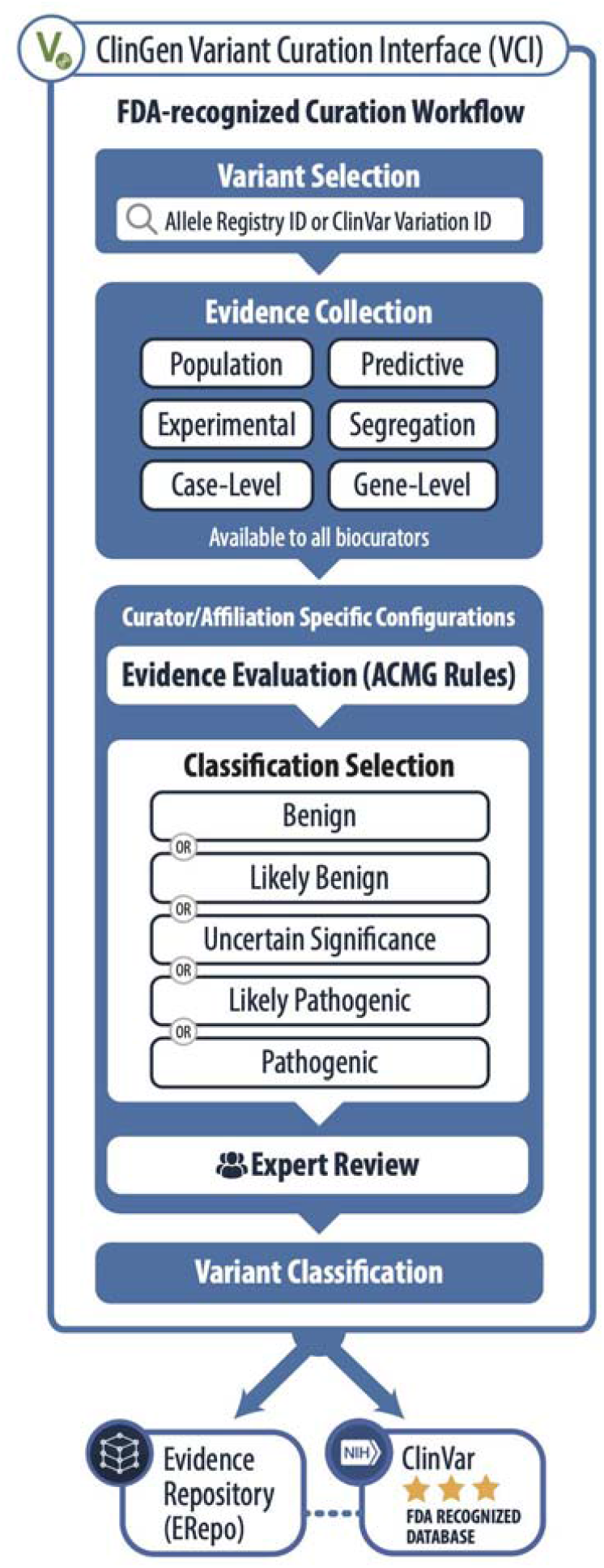
ClinGen FDA-Recognized Variant Curation Process and VCI. Overview of the ClinGen Variant Curation Process using the VCI, an FDA-recognized workflow. Biocurators select a variant, and evaluate evidence that falls into six categories. VCI viewers may view all evidence available for any variant using the VCI. The VCI supports users in making a final pathogenicity classification keeping with the ACMG/AMP guidelines. ClinGen expert panels then disseminate their variant classifications through two community resources: the Evidence Repository (ERepo) and ClinVar.

## Implementation

The VCI curation platform has been developed to facilitate the FDA-recognized ClinGen variant classification process, support transparent evidence review, and provide timely dissemination to the genomics community. Users can curate individually or communally in groups known as affiliations. The VCI programmatically displays relevant data types from external sources (**Table 1)**, and displays evidence identified by other VCI users in an organized user interface enabling an environment to document ACMP/AMP criteria codes.

**Table 1:** VCI Displayed Information Type and Source.

| Information Type | Displayed Data | Data source |
| --- | --- | --- |
| Basic Information | • Variant ID<br>• HGVS terms | ClinGen Allele Registry<sup>26</sup> |
|  | • ClinVar Variation ID<br>• ClinVar Overall interpretation<br>• ClinVar Submitted interpretations<br>• ClinVar Primary transcript<br>• RefSeq transcripts<br>• dbSNP variant ID<br>• Entrez Gene ID | NCBI E-utilities<sup>27</sup> |
|  | • RefSeq transcripts<br>• Ensembl transcripts<br>• Molecular consequences | Ensembl VEP<sup>28</sup> |
|  | • Monarch Disease Ontology (Mondo) human disease term(s) | Ontology Lookup Service<sup>29</sup> |
|  | • Phenotypic abnormality term(s) | Human Phenotype Ontology (HPO)<sup>19</sup> |
| Population | • Allele frequencies<br>○ gnomAD<br>○ ExAC<br>○ Exome Sequencing Project | MyVariant.info<sup>30</sup> |
|  | • Allele frequencies<br>○ PAGE Study | GGV Browser<sup>31</sup> |
|  | • Allele frequencies<br>○ 1000 Genomes | Ensembl VEP<sup>28</sup> |
| Variant Type | • *In silico* predictor scores<br>○ REVEL<br>○ SIFT<br>○ PolyPhen2<br>○ LRT<br>○ MutationTaster<br>○ MutationAssessor<br>○ FATHMM<br>○ PROVEAN<br>○ MetaSVM<br>○ MetaLR<br>○ CADD<br>○ FATHMM-MKL<br>○ fitCons<br>• Conservation analysis scores | MyVariant.info<sup>30</sup> |
|  | ○ phyloP100way<br>○ phyloP30way<br>○ phastCons100way<br>○ phastCons30way<br>○ GERP++<br>○ SiPhy |  |
| Experimental | ● Experimental functional data | ClinGen Functional Data Repository (FDRepo) |
| Gene-Centric | ● Gene symbol | HGNC<sup>32</sup> |
|  | ● ExAC constraint scores<br>● UniProt protein ID<br>● GeneCards gene | MyGene.info<sup>33</sup> |

The core elements of the VCI data model are shown in **Figure 2**. The full data model is stored in Javascript Object Notation (JSON) format with references to data elements. The data model is centered upon a variant classification, with attributes consisting of data and context related to asserting the variant’s pathogenicity. The classification model is based on the combined variant, disease, and mode of inheritance data models. Each variant is evaluated by biocurators against specific evidence types which are reflected in the VCI’s data models, (e.g. population data, experimental data, computational data), as well as literature-based evidence which can be manually added by a biocurator and related to any of the other evidence types. Biocurator-selected evaluations of the evidence criteria provide a computed variant pathogenicity, which can be manually overridden by biocurators based on expert opinion consistent with the ACMG/AMP guidelines.

**Figure 2:**
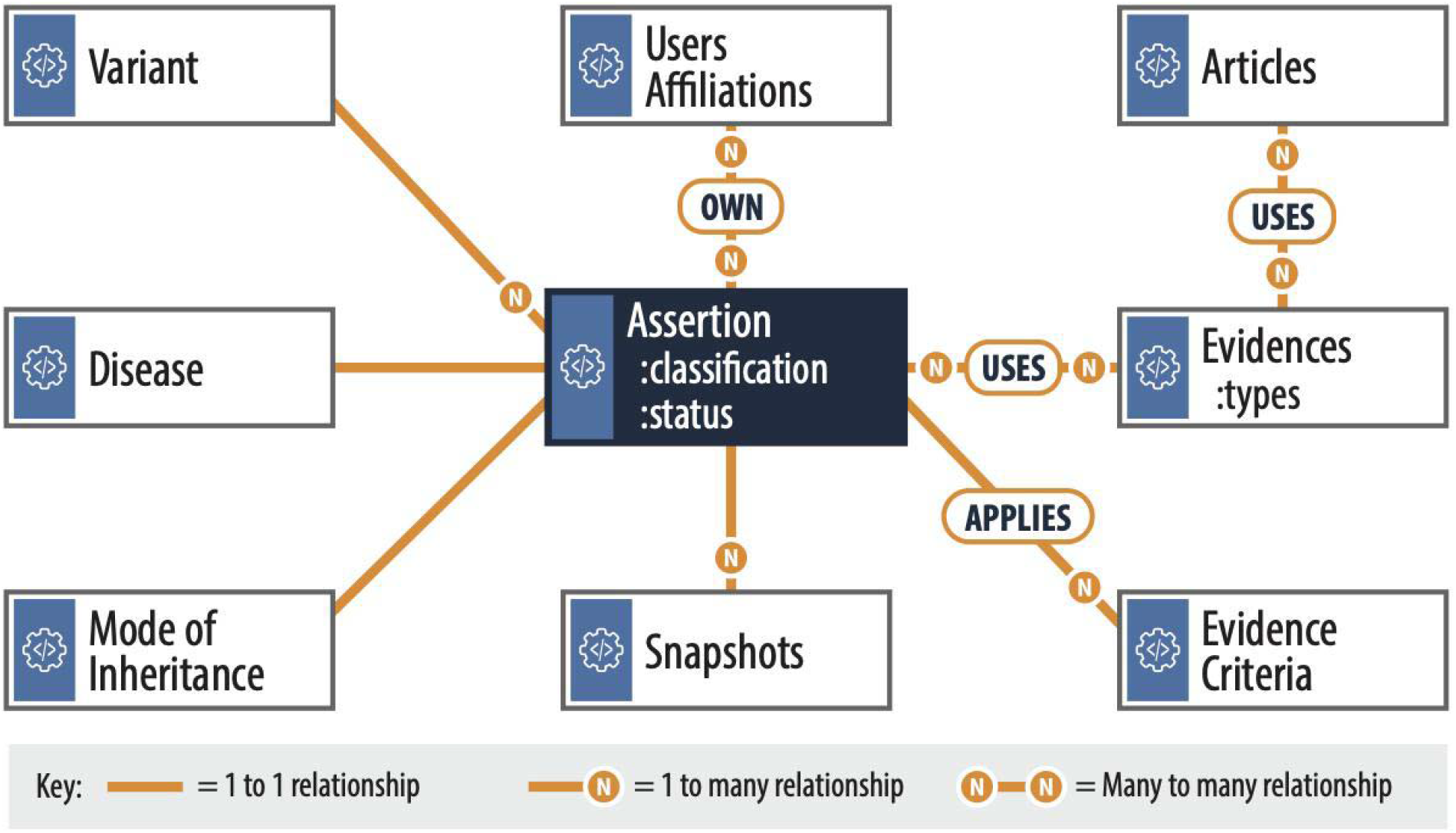
Core VCI Data Model used for storing and retrieving data as JSON documents. The VCI uses a data model centered on a classification (dark blue center box), with relationships to other data models (white boxes). Each assertion is uniquely defined by the Variant, Disease and Mode of Inheritance models, and is owned by a user or affiliation. It has two core attributes a) status indicating its state in the current workflow cycle and b) selected classification (Figure 1). It uses different types of Evidence (see Evidence Collection in Figure 1) and applies an Evidence Criteria to arrive at the selected classification. Each evidence type can use Articles as supplementary evidence. As an interpretation progresses through review statuses (such as review, approved, and published), snapshots of the full data at each review step are created. The relationships between data models are represented here, with 1:1 (solid green lines), 1 to many (N), or many to many (N to N) indicated.

The classification model is designed to support the ClinGen workflow of variant curation, which is an iterative and manual process, with biocurators making criteria evaluations, and expert panels considering and approving final classifications. This is modeled in the platform in two ways, first each variant classification has a status attribute that indicates the most recent status of the classification, (‘In-progress’, ‘Provisional’, ‘Approved’, ‘Published’, ‘New-Provisional’). Secondly, the classification model only stores references to its related models (e.g. variant, disease, evidence, evidence criteria), which also store the most recent information. When a classification has a status of ‘Provisional’, ‘Approved’ or ‘Published’, the snapshot model is used to store an instance (point in time) containing all of the related data. This allows changes to the related data to be identified, while previous data, which may have been used to make a criteria evaluation, are still preserved in the snapshot. Snapshots published to ERepo and ClinVar undergo a transformation to SEPIO (http://dataexchange.clinicalgenome.org/interpretation/index.html).

The VCI is accessed through a web browser where users can perform curation activities including review of imported evidence, entry of evidence gathered from published and unpublished sources, ACMG/AMP criteria application, pathogenicity evaluation, and classification review and approval. The classified variants from ClinGen-approved Variant Curation Expert Panels (VCEPs) are then shared with the ClinGen Evidence Repository (ERepo; https://erepo.clinicalgenome.org/) and the ClinVar database (https://www.ncbi.nlm.nih.gov/clinvar/) to enable peer review and public access.

The software for the VCI is freely and openly available via publicly accessible web pages and a publicly available GitHub repository (https://github.com/ClinGen/clincoded/). The VCI website (https://curation.clinicalgenome.org) is developed as a common interface for both the VCI and the related Gene Curation Interface (GCI), using the same platform and sharing components such as a user database and classification data. User access to the VCI and GCI is available by authenticated login. Login permissions are required in order to document the provenance of evidence added to the interfaces.

Users access the VCI web application via the browser and execute the workflow tasks needed to perform variant curation. The current deployment, VCI v2.0, utilizes cloud-based web development best practices and a “serverless architecture”, which is a cloud development approach where all application resource management and scaling needs are automatically determined and handled by the cloud services. All of the components essential for the application including authentication, gateways to receive and respond to browser requests, microservices, and storage are provided by Amazon Web Services (AWS). This scalable and robust architecture is based on several AWS serverless components shown in **Figure 3**, the role of key components are further described here. The API Gateway handles tens of thousands of requests per second and provides automatic schema validation of data, ensuring data integrity in the VCI. Lambda spawns microservices to store or retrieve data, managing and scaling the computing resources required by the VCI. DynamoDB is a flexible, document-based database that provides constant load-independent performance, supporting the VCI in long-term goals of scaling to large numbers of variant classifications and providing bulk variant curation support, while S3 stores the large VCI database. Additionally, Cognito is used for user management, and Amplify for web content integration with backend microservices. The user interfaces are created using standard JavaScript programming (ReactJS) and they obtain information from the database via an Application Programming Interface (API) using a standard JSON format. For comparison, this continuous integration and deployment provides greater reliability and cost-savings relative to the initial deployment of the VCI v1.0 built following a classical three-tier architecture with a web-frontend component (ReactJS), backend business logic layer (Python and Pyramid), a split persistence layer containing the state and metadata database (PostgreSQL) and search indexes (AWS Elasticsearch).

**Figure 3:**
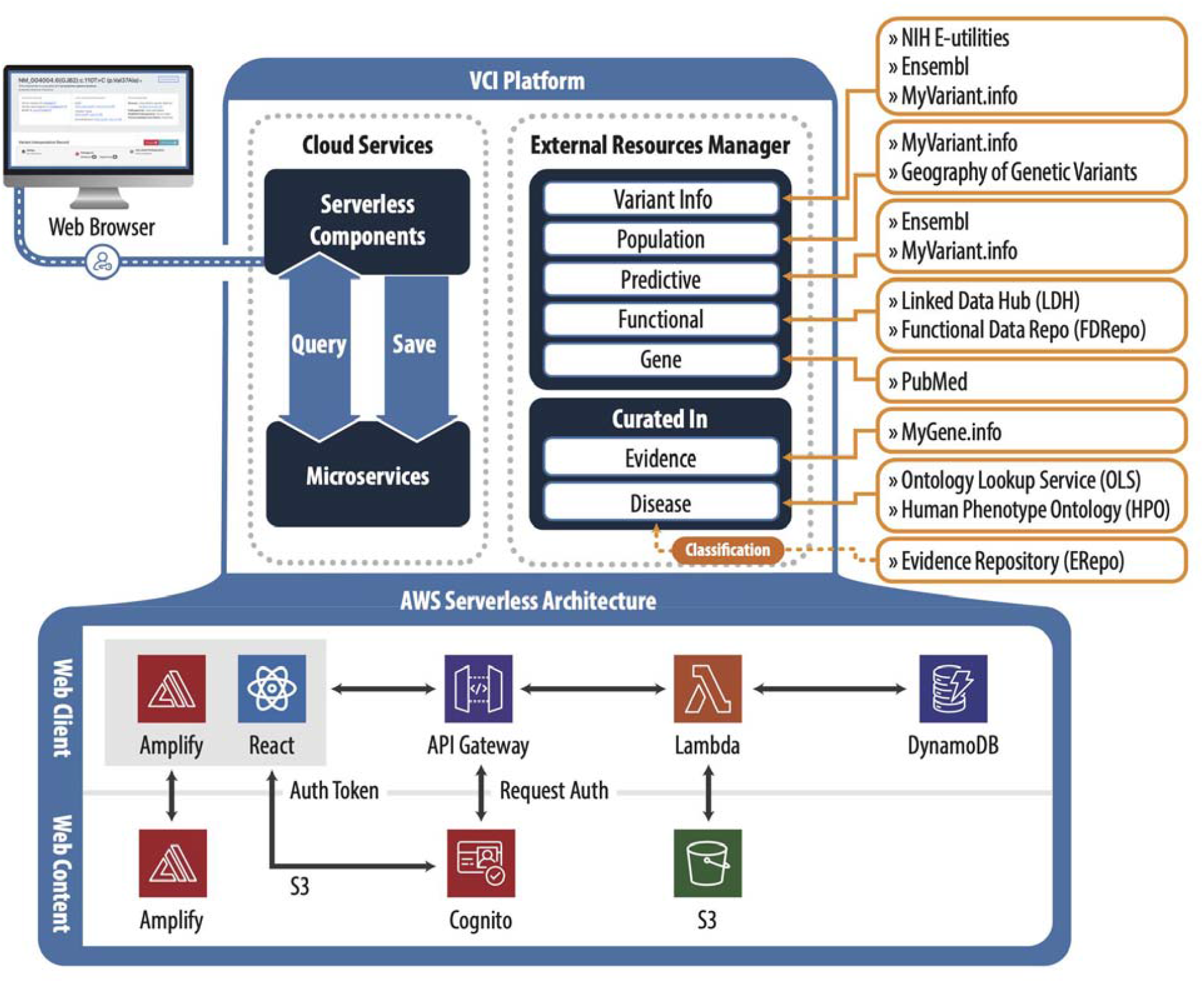
VCI Platform Components Overview Including Schema and Serverless Architecture. The platform is web-browser based based and uses AWS cloud services. An External Resources Manager retrieves population, predictive, functional and other variant and gene-level data from external sources (**Table 1**) via APIs. In addition, the user can add in curated evidence, evaluate against ACMG/AMP guidelines and save the classification for review and approval. The approved classifications are then submitted to the ClinGen Evidence Repository. All data are saved in a database via microservices and can be accessed via queries. The Amazon Cloud Services provide the microservices to store and retrieve data adopting a Serverless Architecture utilizing the following components and services: Amplify, Cognito, API Gateway, Lambda, DynamoDB, S3, and the user-facing web interface is created using React Javascript Library.

Services components include an external resource manager, which is responsible for obtaining data from external sources (**Table 1**) that feed into the variant, gene, disease, population, predictive, functional and gene data models.

Finally, ClinGen provides supplemental resources for VCI users including general information about biocuration, summary videos outlining the concepts and methods behind biocuration, and links to biocuration resources such as ClinGen’s documented standard operating procedures (https://clinicalgenome.org/curation-activities/variant-pathogenicity/).

### Development Process

The VCI software and product development teams worked alongside the ClinGen Variant Curation Interface Task Team to develop the initial platform. This product was designed through a user engagement process and VCI v1.0 was launched for use in September 2016, with new features developed and released monthly. The completely re-architected and updated VCI v2.0 platform was launched in December 2020, and is the current production version.

VCI development continues to evolve with the input of core members of the ClinGen variant curation community that meet twice per month with the VCI development team. This group includes members of the ClinGen Sequence Variant Interpretation (SVI) Working Group, which provides guidance on how to interpret, refine, and standardize the ACMG/AMP guidelines^5–7,11,16^, and members of ClinGen’s Variant Curation Expert Panels (VCEPs). Additional guidance for VCI development comes from the ClinGen Data Access, Protection and Confidentiality (DAPC) Working Group, which reviews tools and data practices in the ClinGen curation ecosystem to ensure that software development efforts are informed by updated data sharing policies. Detailed best practice recommendations for biocurator use of the VCI and associated resources for variant classification are provided in the ClinGen Variant Curation Standard Operating Procedure.

### Variant Identification and Evidence

It is possible to define a variant in several different ways. This promiscuity arises as a result of the availability of multiple transcripts and genomic reference sequences and various ways to describe insertion/deletions. As a result, unambiguous identification of variants is critical to the downstream usability of the curated data. The VCI identifies a variant by either a ClinVar Variation ID^17^ or a ClinGen Allele Registry ID^18^. The variant is then associated with a Mondo Disease Ontology^18^ term and a Human Phenotype Ontology^19^ mode of inheritance term by the biocurator or VCEP. Recognizing that a variant can be curated for more than one potential disease, each user is restricted to one variant classification record per variant. Within the interfaces each variant is titled based on a hierarchy of naming conventions, preferentially using a title based on the MANE Select^20^ transcript when available (**Figure 4A**).

**Figure 4:**
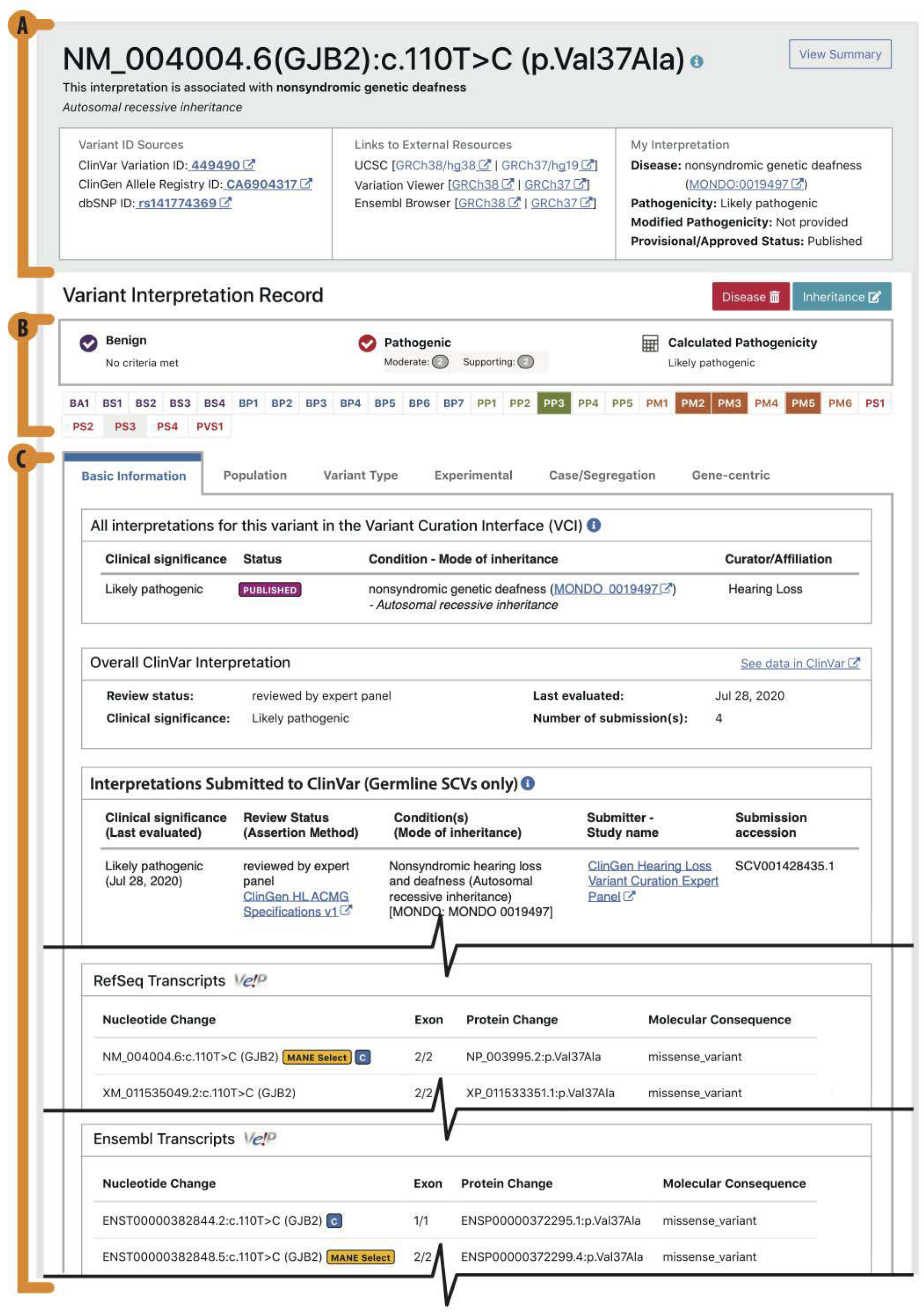
The Basic Information View. The VCI has six tab views that collate and display variant information from external and internal sources to biocurators. **(A)** The top title view is always viewable and shows the variant title, links to the variant in external resources, and key curation information for the record. **(B)** The criteria bar displays evaluated criteria and the calculated pathogenicity. **(C)** The basic information tab displays any curations available for the variant in the VCI and ClinVar, and transcript information from RefSeq and Ensembl.

The VCI aggregates and displays multiple types of evidence about a variant, separated into six tabs structured by data type, providing a rich and structured evidence gathering experience to biocurators, while supporting variant classification in accordance with the ACMG/AMP guidelines. This promotes consistency in terms of the evidence evaluated, application of the ACMG/AMP criteria, and pathogenicity calculations. In keeping with the ClinGen goal to support appropriate community data sharing, all evidence added by users is viewable by any other VCI user. While all evidence is viewable by all users, a user’s evidence evaluations and pathogenicity calculations remain private until the classification record has been finalized (set to ‘Approved’ status), at which point other users can view, but not edit the final classification in the VCI.

### Automated Evidence

The VCI programmatically retrieves and displays many different types of evidence for each variant (**Table 1**). This includes the many possible variant nomenclatures on different transcripts and human genome builds, population frequency data, *in silico* prediction scores, conservation data, gene and protein resources, and all classifications and submissions for the variant currently present in ClinVar or the VCI. When evidence is unavailable for direct display via API, dynamic links to external information sources are embedded within the relevant evidence tab. Relevant external information sources are identified in conjunction with core members of the ClinGen variant curation community described above.

### Manually-Curated Evidence and Structured Data Capture

Users can manually add information relevant to the variant being classified from published articles for any evidence type into the relevant sections in the VCI. Additionally, structured data capture is supported for published functional data, and for published and unpublished case and segregation evidence. Such structured data inputs ensure curation consistency, as they organize and accurately define the information so that it can easily be retrieved, facilitate searching of the captured data, and enable downstream data processing applications such as data mining and machine learning. Two examples of structured data capture within the VCI are outlined below (Functional Data Capture and Case Level Data Capture).

### Functional Data Capture

The ACMG/AMP guidelines require the assessment of well established *in vivo* or *in vitro* functional studies showing ‘no damaging effect’ (BS3) or ‘supportive of damaging effect’ (PS3) on protein function or splicing. We have developed a structured framework with the narrative of 1) method, 2) material, and 3) effect (with or without a quantifier), using standardized terminology from ontologies, in order for users to define the functional data they have derived from published articles in a consistent and reproducible way. We provide users with a standardized template for capturing these structured data which they can then submit to ClinGen’s Functional Data Repository (FDRepo [https://ldh.genome.network/fdr/ui/]); subsequently, these granular functional data for each variant are viewable in the ‘Experimental’ tab in the VCI. Future enhancements will include updating the structured narrative and data fields in accordance with new standards^7^ and augmenting capacity for bulk annotations to be imported from literature annotations or databases of functional evidence including the increasing availability of data from multiplex assays of variant effect^21,22^.

### Case Level Data Capture

Case level and segregation level data are critical to pathogenicity evaluations using the ACMG/AMP guidelines. However, individual case observations have the potential to be linked to individual patient identities if enough ancillary information is also included about the case. It is for this reason that the case-segregation tab of the VCI prompts users to remember the Terms of Use for this tool, which include prohibitions on entering protected health information (PHI) or other sensitive information that could possibly identify an individual data subject, and entering only the minimum necessary information to resolve a particular case. In order to further protect data subjects from the possibility of re-identification (which is also strictly prohibited for users of the VCI as stated in the Terms of Use), individual case-level data are not made publicly available through ClinGen tools except in aggregate. When entering in case observations or pedigree segregation evidence, users are directed to a form that has separate fields to capture each distinct case-level observation or co-segregation events. Then, individual counts for each distinct data type are summed together and analyzed in aggregate along with the same information from other evidence sources (**Figure 5B**).

**Figure 5:**
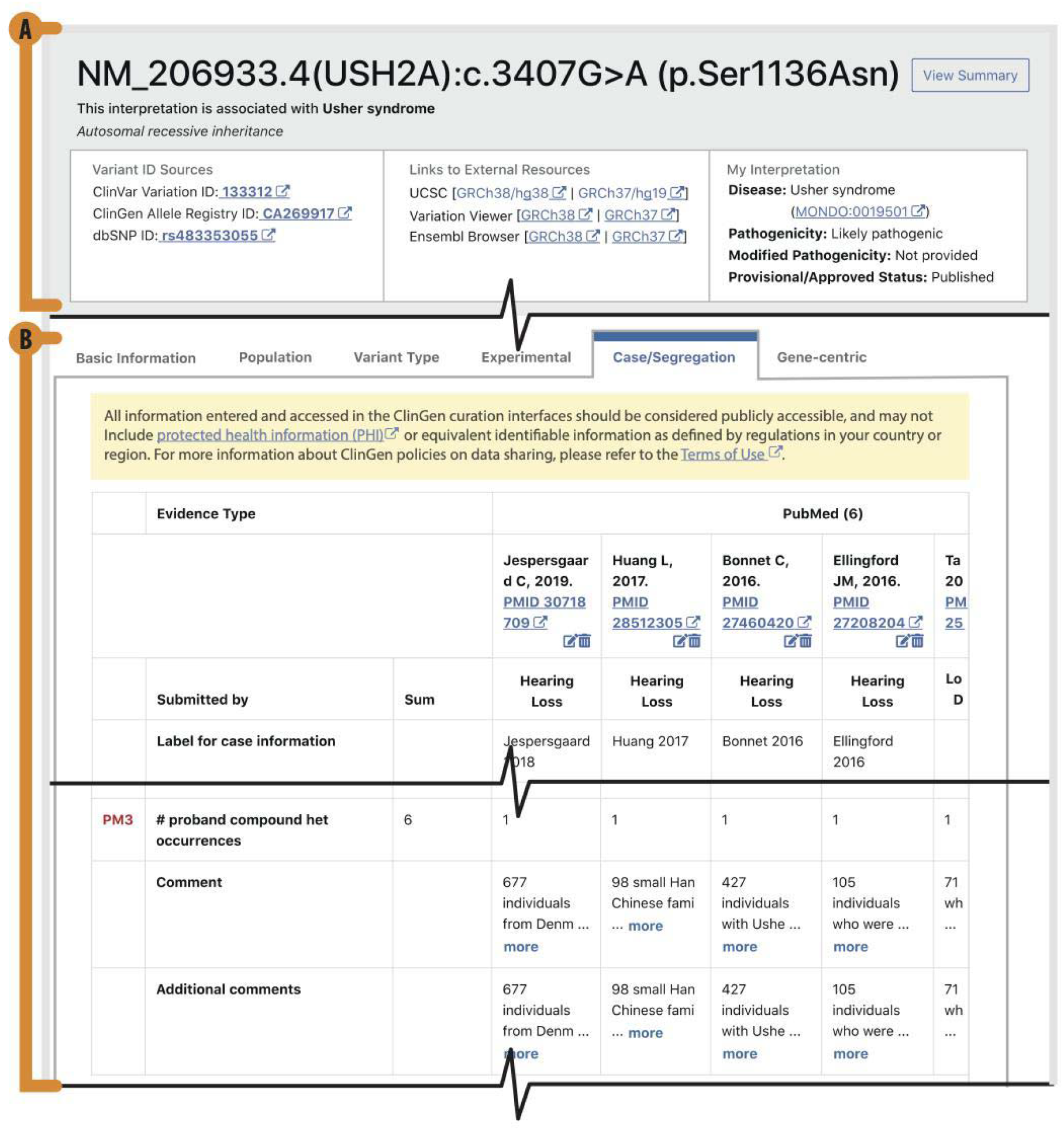
Case and Segregation Evidence Capture. The structured data capture for the case/segregation view in the VCI, including the **(A)** top title view **(B)** Evidence sources are captured and structured so that users can quickly see all sources and the summed individual counts from the pooled evidence for specific ACMG/AMP criteria (shown here is PM3).

### Curation Workflow

ClinGen variant curation through the VCI enables use of the nomenclature, criteria codes and rules defined in the joint 2015 ACMG/AMP guidelines on variant classification. Embedded flexibility is designed to allow biocurators to incorporate modifications and additional guidance produced by ClinGen’s Sequence Variant Interpretation WG as well as disease specifications from ClinGen VCEPs following the FDA-recognized validation process. To further aid this process, the VCI allows groups of users to curate variants as a single entity, known as an ‘affiliation’. VCI affiliations are often ClinGen Variant Curation Expert Panels (VCEPs)^23,24^; however, any group of users who wish to curate variants together (e.g. a clinical or research laboratory) may form an affiliation. Once a VCI user initiates a classification it belongs to that individual or affiliation, and can only be edited by them.

The ACMG/AMP guidelines provide a set of criteria to be considered when classifying a variant. The VCI is designed to help users evaluate the applicability of these criteria in an efficient and structured way (**Figure 1**). The VCI groups criteria by evidence types; displaying both the relevant criteria and any related evidence. These groups are: 1) Population (known variant allele frequencies), 2) Variant Type (predicted impact of the variant on the gene product), 3) Experimental (functional assay data), and 4) Case/Segregation (relevant observations of the variant). The VCI also groups together gene-focused resource links, and basic information, displaying ClinVar and VCI curations for the variant as well as the molecular consequence of the variant on all known transcripts (**Figure 4C** and **Table 1**).

For each ACMG/AMP criteria, users are provided a description of the guideline in the VCI. The user is able to view, add and evaluate the relevant evidence, and then set their criteria evaluation and write an explanation. As some criteria are applicable at different pathogenicity strengths, users can choose the appropriate strength from a pulldown list containing only the appropriate strength options for that criterion. For instance, the available options provided for evaluating PP1 are: Not Evaluated, Met, Not Met, PP1_Moderate, and PP1_Strong. As a user saves their evaluations, a criteria bar (**Figure 4B**) in the header of the interface keeps track of their progress by indicating which criteria have been ‘Met’ (solid color background with white criteria code), ‘Not Met’ (grey background with colored criteria code), or remain ‘Not Evaluated’ (white background with colored criteria code). If a user scrolls over individual criteria codes in this bar they will see a description for each criteria and they can click on individual criteria codes to link to the pertinent section in the VCI. Additionally, a progress bar shows the number of criteria met according to the strength of the evaluation and whether they are ‘Benign’ or ‘Pathogenic’ and also automatically calculates the pathogenicity each time an evaluation is saved or updated. At any time a user can view an ‘Evaluation Summary’ that summarizes all their evaluated evidence. If a criterion code is not evaluated then it would not be considered in the calculation of a predicted classification. Once a biocurator is satisfied they have reviewed all pertinent evidence, and evaluated all relevant criteria, they can save their classification as ‘Provisional’. This generates a PDF version of the ‘Evaluation Summary’ that can be distributed among the VCEP membership domain experts to aid in their review process. Upon satisfactory completion of the review process a final classification can be saved as ‘Approved’, at which point the ‘Evaluation Summary’ can now be viewed by all VCI users.

### FDA Recognition and Data Dissemination

The VCI generates an output file of the final variant pathogenicity classification in an auto-generated format compatible with ClinVar submission specifications. This is intended to facilitate timely dissemination of variant classifications to the genomics community via ClinVar and is a requirement for ClinGen VCEPs. The ultimate goal is to support fully automated, API-based ClinVar submission through the VCI once ClinVar provides support for API-based submission. Once submitted, a ‘submission to ClinVar’ (SCV) identifier is obtained and can be viewed in the VCI variant record.

The ClinGen variant curation process was recognized by the FDA in December 2018^25^, and is followed by all ClinGen VCEPs. The evidence curation data and pathogenicity classifications generated within the VCI by ClinGen VCEPs are therefore considered to be valid scientific evidence that can be used to streamline the test development and validation processes. As such, additional steps and requirements apply to the information specifically generated through the ClinGen VCEP variant curation and classification process. Specifically, all the evidence that has been curated and evaluated, along with provenance should be made publicly available and easily accessible. With this in mind the VCI saves all evidence that is evaluated by its users. In addition, upon final approval of a classification from a ClinGen VCEP, the VCI facilitates data flow to the ClinGen Evidence Repository (ERepo), where the finalized classifications and associated evidence evaluation is published. Importantly, the VCEP generated variant record in the ERepo includes comments for specific codes enabling in-depth, and transparent data for peer review. (**Figure 1**). These publicly accessible displays of the final ClinGen VCEP variant classifications are accessed via the ERepo API at https://erepo.clinicalgenome.org/evrepo/.

### Current status and users

VCI currently has over 1,100 registered users, two-thirds of whom are members of ClinGen VCEPs (**Figure 6**). The VCI is publicly-accessible (with registration) for variant curation. When curating together as an affiliation, all members can view and edit all information added by anyone in that affiliation. This popular feature is currently used by 79 registered affiliations, most of which represent official ClinGen VCEPs but also include other affiliations of ClinGen members (e.g. institution-specific biocurator teams) and groups unrelated to ClinGen (e.g. clinical and research laboratories).

**Figure 6:**
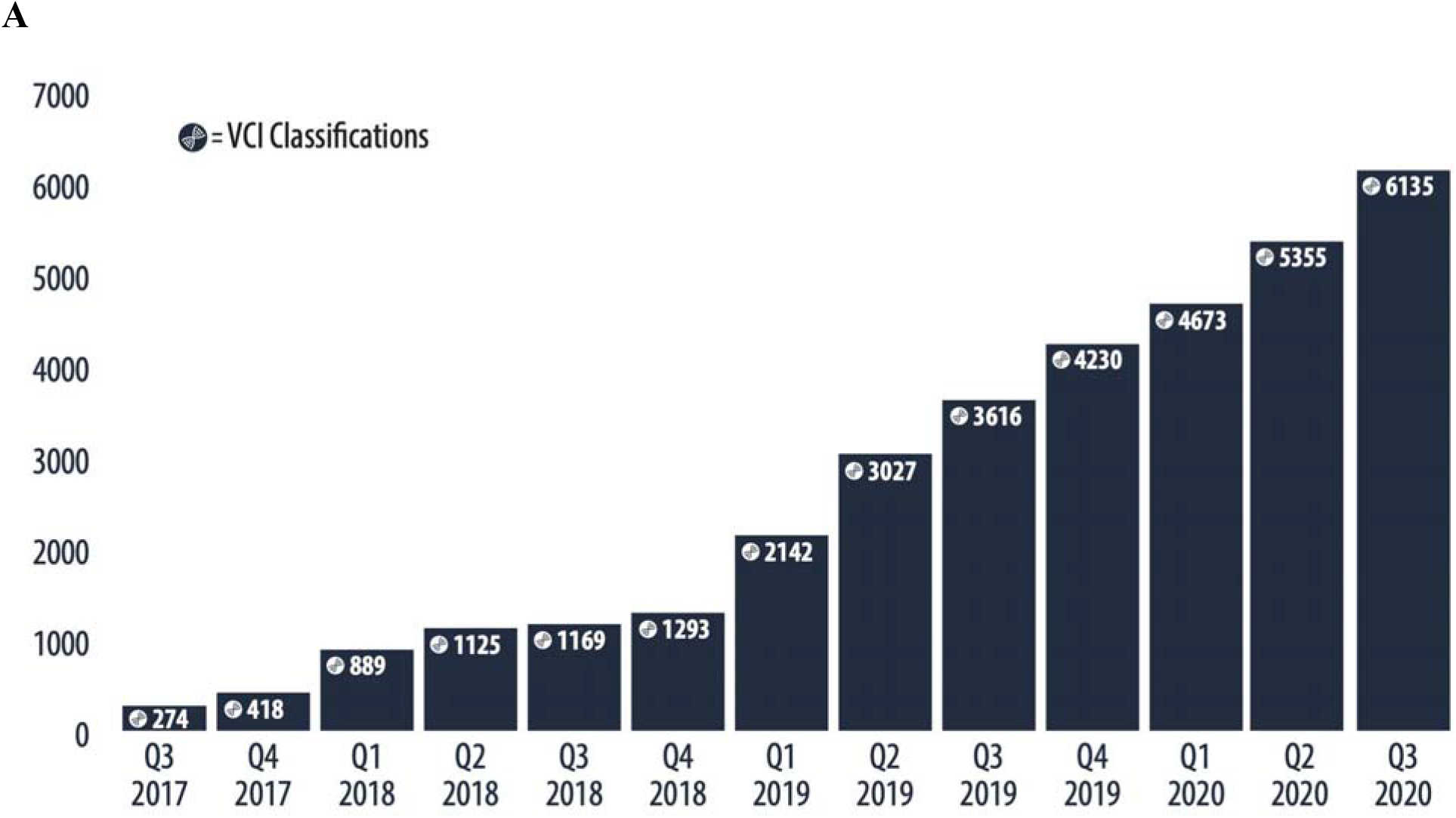
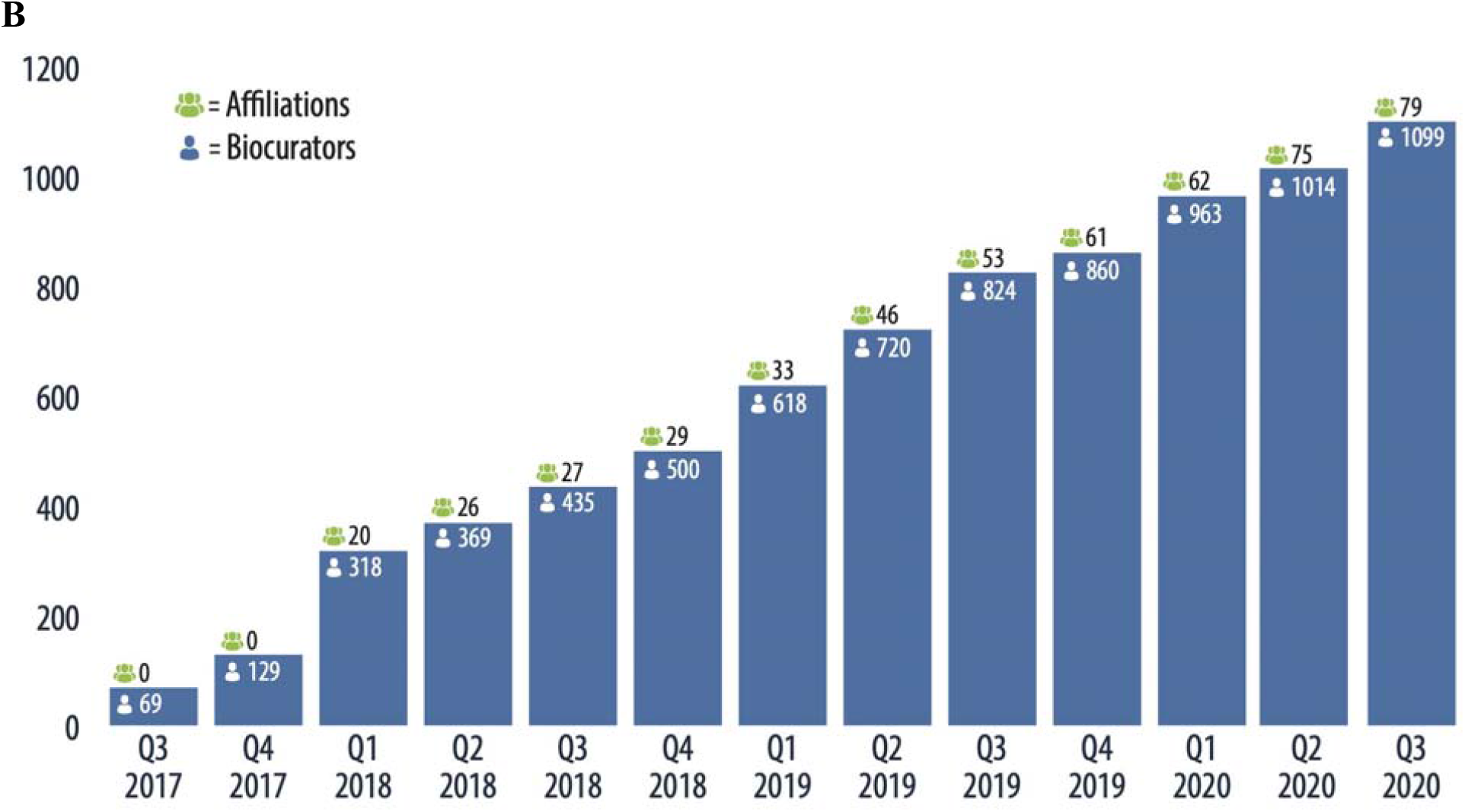
VCI Platform growth over time. **(A)** Number of curated variant classifications performed in the VCI over time. **(B)** The number of biocurators and biocurator affiliations accumulated over time are noted at the top of each bar.

## Discussion

Here we present the development of a genetic variant biocuration platform for health care providers, researchers and the medical genetics community to determine which gene variants are causal for a disease. The VCI supports the FDA-recognized ClinGen variant curation process and combines clinical, genetic, population, and functional evidence with expert review to classify variants into ACMG/AMP 2015 variant classification guideline categories^1^. Primary features of the VCI include the ability to 1) curate individually or in groups, 2) associate pertinent evidence with variant classifications, 3) allow users to assess evidence per variant curation disease/gene-specific protocols, 4) enable users to save provisional records, 5) support an expert review process of curated evidence, and 6) automatically publish classifications and underlying criteria assessments to the ERepo

Future VCI improvements will focus on enhancing scale, workflow, throughput, and support ongoing compliance with FDA recognition of the ClinGen Variant Curation Expert Panels through the FDA Human Variant Database program. The current VCI v2.0 platform has over 6,300 variant classifications in different curation stages, and our modernized architecture is able to scale to support over 1 million future classifications. We plan to enhance workflow usability and curation efficiency by making the platform more proactive with: 1) task management (supporting assigning of variant classification records to users) and action items (alerts to users), 2) support for bulk variant curation workflows, 3) automatically bring in additional variant evidence data and monitor major data changes via a streaming service so curations can be updated as needed, and 4) provide customized curation experiences based on VCEP specifications. To ensure compliance with FDA requirements for the auditability of ClinGen variant curation process, the VCI database maintains a complete audit trail of all saved curation actions. We will further support FDA compliance with 1) additional traceability, 2) permanent archiving, 3) regular knowledge updating through literature and database monitoring, and 4) update alerts provided to curation teams.

The VCI provides needed software infrastructure and a comprehensive curation platform necessary for supporting variant classification, a critical step in the use of genomics in medicine. This global open source platform aids individual biocurators and teams of collaborating biocurators in performing the complex task of variant curation in an efficient workflow to enforce rigor and quality in variant classification ultimately contributing to scientific advancement and informing health care management.

## Data Availability

All code for the VCI including front-end, back-end, database schemas, analysis pipelines, and user interfaces are freely available under MIT Open Source licenses via the GitHub repositories (https://github.com/ClinGen/ and https://github.com/ClinGen/clincoded/).

## Acknowledgements

All authors contributed to the project design. C.G.P., M.W.W., R.M., and H.A.C. drafted the initial version of the manuscript. All authors contributed to and approved the final version of the manuscript. We would like to thank all of the members of the Clinical Genome Resource consortium, especially the core members attending bimonthly VCI feedback meetings for their continual feedback.

## References

1. Richards, S. et al. Standards and guidelines for the interpretation of sequence variants: a joint consensus recommendation of the American College of Medical Genetics and Genomics and the Association for Molecular Pathology. Genet. Med. 17, 405–424 (2015).

2. Niehaus, A. et al. A survey assessing adoption of the ACMG-AMP guidelines for interpreting sequence variants and identification of areas for continued improvement. Genetics in medicine: official journal of the American College of Medical Genetics vol. 21 1699–1701 (2019).

3. Rehm, H. L. et al. ClinGen--the Clinical Genome Resource. N. Engl. J. Med. 372, 2235– 2242 (2015).

4. Amendola, L. M. et al. Performance of ACMG-AMP Variant-Interpretation Guidelines among Nine Laboratories in the Clinical Sequencing Exploratory Research Consortium. Am. J. Hum. Genet. 98, 1067–1076 (2016).

5. Abou Tayoun, A. N. et al. Recommendations for interpreting the loss of function PVS1 ACMG/AMP variant criterion. Hum. Mutat. 39, 1517–1524 (2018).

6. Biesecker, L. G., Harrison, S. M. & ClinGen Sequence Variant Interpretation Working Group. The ACMG/AMP reputable source criteria for the interpretation of sequence variants. Genetics in medicine: official journal of the American College of Medical Genetics vol. 20 1687–1688 (2018).

7. Brnich, S. E. et al. Recommendations for application of the functional evidence PS3/BS3 criterion using the ACMG/AMP sequence variant interpretation framework. Genome Med. 12, 3 (2019).

8. Luo, X. et al. ClinGen Myeloid Malignancy Variant Curation Expert Panel recommendations for germline RUNX1 variants. Blood Advances vol. 3 2962–2979 (2019).

9. Lee, K. et al. Specifications of the ACMG/AMP variant curation guidelines for the analysis of germline CDH1 sequence variants. Hum. Mutat. 39, 1553–1568 (2018).

10. Oza, A. M. et al. Expert specification of the ACMG/AMP variant interpretation guidelines for genetic hearing loss. Hum. Mutat. 39, 1593–1613 (2018).

11. Ghosh, R. et al. Updated recommendation for the benign stand-alone ACMG/AMP criterion. Hum. Mutat. 39, 1525–1530 (2018).

12. Mester, J. L. et al. Gene-specific criteria for PTEN variant curation: Recommendations from the ClinGen PTEN Expert Panel. Hum. Mutat. 39, 1581–1592 (2018).

13. Zastrow, D. B. et al. Unique aspects of sequence variant interpretation for inborn errors of metabolism (IEM): The ClinGen IEM Working Group and the Phenylalanine Hydroxylase Gene. Human Mutation vol. 39 1569–1580 (2018).

14. Gelb, B. D. et al. ClinGen’s RASopathy Expert Panel consensus methods for variant interpretation. Genetics in Medicine vol. 20 1334–1345 (2018).

15. Kelly, M. A. et al. Adaptation and validation of the ACMG/AMP variant classification framework for MYH7-associated inherited cardiomyopathies: recommendations by ClinGen’s Inherited Cardiomyopathy Expert Panel. Genetics in Medicine vol. 20 351–359 (2018).

16. Tavtigian, S. V. et al. Modeling the ACMG/AMP variant classification guidelines as a Bayesian classification framework. Genet. Med. 20, 1054–1060 (2018).

17. Landrum, M. J. et al. ClinVar: public archive of relationships among sequence variation and human phenotype. Nucleic Acids Res. 42, D980–5 (2014).

18. Shefchek, K. A. et al. The Monarch Initiative in 2019: an integrative data and analytic platform connecting phenotypes to genotypes across species. Nucleic Acids Res. 48, D704– D715 (2020).

19. Köhler, S. et al. Expansion of the Human Phenotype Ontology (HPO) knowledge base and resources. Nucleic Acids Res. 47, D1018–D1027 (2019).

20. Matched Annotation from NCBI and EMBL-EBI (MANE). https://www.ncbi.nlm.nih.gov/refseq/MANE/.

21. Esposito, D. et al. MaveDB: an open-source platform to distribute and interpret data from multiplexed assays of variant effect. Genome Biol. 20, 223 (2019).

22. Gelman, H. et al. Recommendations for the collection and use of multiplexed functional data for clinical variant interpretation. Genome Med. 11, 85 (2019).

23. Harrison, S. M., Biesecker, L. G. & Rehm, H. L. Overview of Specifications to the ACMG/AMP Variant Interpretation Guidelines. Curr. Protoc. Hum. Genet. 103, e93 (2019).

24. Rivera-Muñoz, E. A. et al. ClinGen Variant Curation Expert Panel experiences and standardized processes for disease and gene-level specification of the ACMG/AMP guidelines for sequence variant interpretation. Hum. Mutat. 39, 1614–1622 (2018).

25. Office of the Commissioner. FDA takes new action to advance the development of reliable and beneficial genetic tests that can improve patient care. https://www.fda.gov/news-events/press-announcements/fda-takes-new-action-advance-development-reliable-and-beneficial-genetic-tests-can-improve-patient (2018).

26. Pawliczek, P. et al. ClinGen Allele Registry links information about genetic variants. Hum. Mutat. 39, 1690–1701 (2018).

27. NCBI Resource Coordinators. Database resources of the National Center for Biotechnology Information. Nucleic Acids Res. 46, D8–D13 (2018).

28. McLaren, W. et al. The Ensembl Variant Effect Predictor. Genome Biol. 17, 122 (2016).

29. Madeira, F. et al. The EMBL-EBI search and sequence analysis tools APIs in 2019. Nucleic Acids Res. 47, W636–W641 (2019).

30. Xin, J. et al. High-performance web services for querying gene and variant annotation. Genome Biol. 17, 91 (2016).

31. Marcus, J. H. & Novembre, J. Visualizing the geography of genetic variants. Bioinformatics 33, 594–595 (2017).

32. Braschi, B. et al. Genenames.org: the HGNC and VGNC resources in 2019. Nucleic Acids Res. 47, D786–D792 (2019).

33. Wu, C., Macleod, I. & Su, A. I. BioGPS and MyGene.info: organizing online, gene-centric information. Nucleic Acids Res. 41, D561–5 (2013).

